# Geo-epidemiological risk stratification to optimize malaria control interventions, case of Mali

**DOI:** 10.1101/2025.05.30.25328661

**Authors:** Mady Cissoko, Mahamadou Magassa, Ibrahim A. Cissé, Seybou Coulibaly, Daouda S. Samaké, Vincent Sanogo, Mamady Koné, Sylla Thiam, Tako Ballo, Samba Diarra, Seydou Fomba, Aïssata Koné, Jean Gaudart, Issaka Sagara

## Abstract

**Introduction:** Malaria is a public health threat in Mali, with high morbidity (37%) and mortality (25%) rates. Malaria risk stratification is needed to identify different transmission zones and prioritize the interventions, especially in resource-limited contexts. For the new National Strategic Plan, we updated stratification with World Health Organization (WHO) recommendation. This study presents the selection of interventions based on stratification.

**Method:** Data collection covered all the 75 Health Districts (HDs) in the country for the period 2018-2022. This was further supplemented by national survey data on parasitology and entomology in Mali. To estimate the adjusted incidence, the analysis considered health data reporting, malaria diagnostic positivity and health facility attendance rates at health district level to malaria cases estimated. Mixed interventions were defined based on adjusted incidence, prevalence, seasonality, vector resistance to insecticides and parasite distribution by Health Districts or region according to data available.

**Results:** Four strata have been defined according to the 2017 WHO Malaria Elimination Framework. Most HDs (54) fall within the highest and moderate transmission areas, covering 84% of the population (about 18 million) located in the southern and central regions. A total of 19 intervention packages were selected and adapted. We identified 61 out of 75 eligible HDs for the LLINs mass distribution campaign, 57 HDs for SMC, 67 for IPTp including community IPTp and 19 HD for vaccination.

**Conclusion:** A half of Malian population live in high-risk areas. These districts require a continuous intensification of interventions. Malaria stratification was critical for strategic planning and appropriate deployment of malaria control interventions in Mali.

## Introduction

Malaria remains a significant global health threat, with sub-Saharan Africa bearing an unequal proportion of the global burden. In 2023, the World Health Organization (WHO) reported approximately 263 million malaria cases worldwide, resulting in around 597,000 deaths [1]. In Mali, the situation is similar. Malaria is a major health concern and a national priority. The National Health Information System (SNIS) indicates that malaria was the leading cause of medical consultations and deaths in 2023, accounting for a morbidity of 37 % of cases and 25 % of mortality at the national level. Mali is one of the 11 countries in the high burden high impact approach, accounting for 3.1% of the malaria burden [2]. In 2023, the NMCP recorded 3,771,426 clinical cases, including 1,197,864 severe cases and 1,498 deaths, resulting in hospital fatality rate of 1.67‰ [3].

*Plasmodium falciparum (Pf)*, which constitutes 97 percent of malaria cases and most severe malaria cases and deaths, is the primary parasite in Mali. In addition, *Plasmodium vivax (Pv)*, has been reported in the Menaka and Kidal regions (northern Mali). It is the second most prevalent parasite, accounting for 4-21 % of all cases in these areas. Other studies had reported the presence of *P. vivax*. Monitoring is ongoing. *Plasmodium ovale* and *Plasmodium vivax* are less common [4]. The primary vectors for these parasites are mosquitoes of the *Anopheles* genus, more specifically the *Anopheles gambiae* complex [5].

Malaria remains a major health concern and a national priority for Mali. Over the past decade, the National Malaria Control Program (NMCP) has intensified efforts to implement malaria interventions to reduce morbidity and mortality. Key strategies included the distribution of long-lasting insecticidal net (LLINs) through universal coverage campaigns, as well as routine distributions to pregnant women during prenatal visits and to children during measles vaccinations (NSP). The first stratification in 2021 [6] adapted the type of LLIN to resistance in certain health districts [7,8]. The 2023 ITN mass distribution campaign reduced malaria incidence by 10 %. Additionally, Indoor residual spraying (IRS) has been used in specific districts based on transmission patterns [9]. The program has also expanded other highly effective interventions, including rapid diagnostic tests (RDTs) and microscopy, artemisinin-based combination therapy (ACT) for the treatment for uncomplicated malaria case, injectable artesunate/artemether for severe malaria, intermittent preventive treatment with Sulfadoxine-Pyrimethamine (SP) for pregnancy women and the Community Health Worker (CHW) program. There were no changes to these procedures in the implementation of the first stratification [6]. For the Seasonal Malaria Chemoprevention (SMC) in children, the number of cycles has been adapted to the duration of the high transmission season, and the start of the SMC campaign [6,10]. This adaptation of the SMC reduced the operational cost of the campaign by reducing the number of health districts in the north, where the rainy season is short, to 3 cycles. Strengthened epidemiological surveillance has played a crucial role in monitoring and responses effort [11]. As results, malaria cases had significantly reduced, with the prevalence of malaria in children under five dropping from 47% in 2012 to 19% in 2021 [12].

To meet Mali’s goal of a 30 % reduction in malaria incidence and mortality by 2028 [13], it is crucial to ensure universal coverage of key interventions while tailoring strategies to district-specific needs, especially in resource-limited contexts. Stratification will be continued, drawing on the lessons learned from the first experience. It will be used as a basic planning tool.

To support the development of Mali’s National Strategic Plan (NSP) for malaria control (2024-2028) and finalize the concept note for Global Fund funding [6], we conducted a review of data on malaria seasonality, infant and child mortality rates, the prevalence and distribution of parasitic species, vector resistance to insecticides, logistics, security conditions, and funding available. The stratification was updated in 2023. This study presents the selection of interventions based on stratification risk issued from routine activities and health data reporting in Mali.

## Materials and Methods

### Study setting

This study was carried out in Mali, a country covering 1,241,238 [14] square kilometers with an estimated population of 21,9 million and a growth rate of 2.77% (RGPH 2022). Mali experiences rainfall variation: annually precipitation ranges from under 200 mm in desert areas to over 1,100 mm in the pre-Guinean zone. In the southern and central regions, the rainy season begins between April and June, peaking in August, while northern regions have a considerably lower rainy season starting in July or August [10] and typically lasts no more than three months [15].

Mali is organized into 19 regions and the District of Bamako, further divided into 75 health districts and approximately 1,605 community health centers, some of which affected by insecurity. To improve healthcare access in these difficult areas, the Ministry of Health and some partners have implemented the Community Health Worker (CHW) program that provides integrated health services, including diagnosis and treatment for uncomplicated malaria. As for 2023, there were about 3,327 CHWs who diagnosed 8% of the total malaria cases [16,17].

### Study design

We conducted an analysis of data from 75 health districts using annual malaria cases from January 2018 to December 2022, along with environmental variables, parasitological data, and main malaria vector distributions. For the analysis, mixed approaches (qualitative and quantitative) were used, namely descriptive, geospatial and group discussion based on the experiences of the actors [18].

### Data collection, description and sources

#### Routine and population data

The main data source was routine malaria surveillance information collected from the MOH’s Health Management Information System (HMIS) 2018-2022, hosted on the District Health Information Software 2 (DHIS2). The DHIS2 platform aggregates data from public health facilities, referral hospitals and private clinics. The dataset included the number of suspected, presumed, tested and confirmed cases by RDTs or microscopy. These data are reported monthly and categorized by age groups (under 5 years vs 5 years and older). Other Information were also extracted: reporting rates, positivity rates, number of CHWs per health district, total population, population within a 5km radius, and health facilities attendance rates. Additionally, we analyzed data on intermittent preventive treatment in pregnancy (IPTp) for 2022 specifically covering pregnant women with at least 3 doses of Sulfadoxine Pyrimethamine (SP). To estimate reporting completeness, we compared the number of monthly facilities reports received to the expected number, which is 12 monthly reports.

#### Environmental data

The daily rainfall data from remote sensing was used. These data were collected from the Climate Hazards Group InfraRed Precipitation with Station Data (CHIRPS) via the Google Earth engine interface (0.05° resolution) at the level of health district [19]. The data was collected during 2022.

#### Parasitological and entomological data

Parasitological data were collected from scientific studies conducted in 2021 to describe parasitic species and resistance [20].

*Plasmodium falciparum* (*Pf*) and *Plasmodium vivax* (*Pv*) malaria were collected from scientific studies.

The distribution of vector species and their resistance to insecticides was also collected in 2021 through entomological surveillance programs, and tested using bioassays in line with WHO recommendations [21].

#### Data quality

The quality of the monitoring data was acceptable, with a timeliness rate of 98. The rate of testing of suspected cases is estimated at 99%.

#### Data analysis

We used health districts as analysis units. Data from DHIS2 required no additional processing, as the incidence rates per health district were directly used. To estimate malaria transmission in each health district, we applied the thresholds from the WHO Stratification guidelines, which was approved by the national Technical Working Group (TWG). The same criteria were used for the 2020 stratification [6]. Malaria incidence cases per 1,000 inhabitants were calculated for health districts. Malaria cases are reported as crude incidence and expressed as the number of confirmed cases per 1,000 people in the population. For the missing data, the proposed adjustment considered the data not reported.

### Geographic accessibility to healthcare

The issue of geographic accessibility to health facilities was evaluated using two indicators: the proportion of the population beyond a 5 km radius from a health facility, and the population served per each CHW. These indicators were mapped.

The proportion of the population located more than 5 km from a health facility was calculated by dividing this population by the total population.

The population served by each CHW was calculated by dividing the population located more than 5 km from a health facility by the total number of CHWs.

### Rainfall

The daily rainfall data were aggregated to obtain the mean annual rainfall for each health district from 2019 to 2022. This annual rainfall was mapped.

### Stratification

To estimate malaria transmission in each health district, we applied the thresholds from the WHO framework for malaria elimination (see below). The same criteria were used for the 2020 stratification exercise [11,22]. Malaria incidence cases per 1,000 persons year were calculated for each health district. Malaria cases are reported as crude incidence and expressed as the number of confirmed cases per 1,000 people in the population.

Health districts with less than 100 cases per 1,000 person-years or Pf/Pv prevalence between 0% and 1% were classified as "Very Low Transmission Zones". Districts with incidence rates between 100 and 250 cases per 1,000 person-years or *Pf/Pv* prevalence between 1% and 10% were classified as "Low Transmission Zones". Districts with incidence rates between 251 and 450 cases per 1,000 person-years or *Pf/Pv* prevalence between 10% and 35% were classified "Moderate Transmission Zones". And districts with incidence rates above 450 cases per 1,000 person-years or *Pf/Pv* prevalence of 35% or more were classified as "High Transmission Zones" [22].

Initially, each health district was assigned to one of four strata based on the WHO Stratification guidelines.

In the second step, we performed three adjustments: firstly, to include presumed cases that would be likely positive based on the positive rates; secondly, to address malaria cases that were not reported in the national system; and, lastly, to consider the health service utilization rate or the fraction seeking treatment as follows:

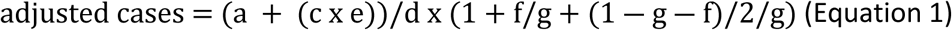

a. *is malaria cases confirmed in the public sector*
b. *is suspected cases tested*
c. *is presumed cases (not tested but treated as malaria)*
d. *is reporting completeness*
e. *is test positivity rate (malaria positive fraction) = a/b*
f. *is the fraction seeking treatment in the private sector*
g. *is the fraction seeking treatment in the public sector or rate of healthcare use in health facilities* *Factor to adjust for those not seeking treatment: (1-g-f)* *Cases in the public sector: (a + (c x e))/d* *Cases in the private sector: (a + (c x e))/d x f/g* [23].

This adjustment method for malaria cases proposes proportion of seeking health care. This indicator is available at regional level. Given the variation between health districts within a region, we replaced it with the attendance rate. By modifying the WHO method for estimating cases, we carried out a sensitivity test. Thus, we determined the difference between the WHO method and the actual method with the attendance rates by health district.

A sensitivity analysis was performed between the local method based on the rate of demand for health care and the one of the WHO based on the proportion of health care seeking. For this, the variation between the number of crude cases (reported) and the cases calculated by the two methods was calculated. The curves of these differences variations were presented, and a Kolmogorov-Smirnov test was performed to evaluate the variance between the two variances.

### Targeted Interventions

Finaly, Interventions per strata were determined in accordance with the WHO’s Global Technical Strategy 2016-2030, the WHO High Burden High Impact initiative, national malaria control policies, and guidelines for malaria prevention and case management. Intervention packages and potential impact on the burden of malaria in each district were discussed and approved by the NMCP, Technique Workshop Group (TWG) and the different partners. Selection of interventions was guided by risk factors strata, seasonality of transmission, infant and child mortality, prevalence and distribution of parasite species, vector resistance to insecticides, logistical considerations, security conditions, and funding availability. The IPTp rate was used to prioritize eligibility health districts with low coverage rate, which will require community-based IPTp(c-IPTp). To improve the coverage of ITP, c-IPTp was selected for health districts with high and moderate transmission, with CHWs and annual coverage below the national average of 49.5%.

The package of interventions and the potential impact on the burden of malaria in each health district were the subject of discussion and approval by the NMCP and the various partners.

### Software analysis

Data analysis used QGIS.org software version 3.10.13 (2020). QGIS Geographic Information System, Open-Source Geospatial Foundation Project. http://qgis.org ».

### Ethics approval consideration

The study was approved by the National Malaria Control Program (042/MSDS-SG/NMCP, 14 January 2023), in accordance with ethics and medical research in Mali. Data was collected through routine disease surveillance.

## Results

### Geographic accessibility to healthcare

This section focused on geographical accessibility and where the deployment of CHWs is highly needed. The proportion of the population beyond a 5 km radius from a health facility, and population served per each CHW were used to prioritize the deployment of CHWs as part of mix of interventions, especially in remote or isolated locations.

Results indicate that over half of the health districts had a large population living beyond 5 km from a health facility (see Fig 1).

**Fig 1:**
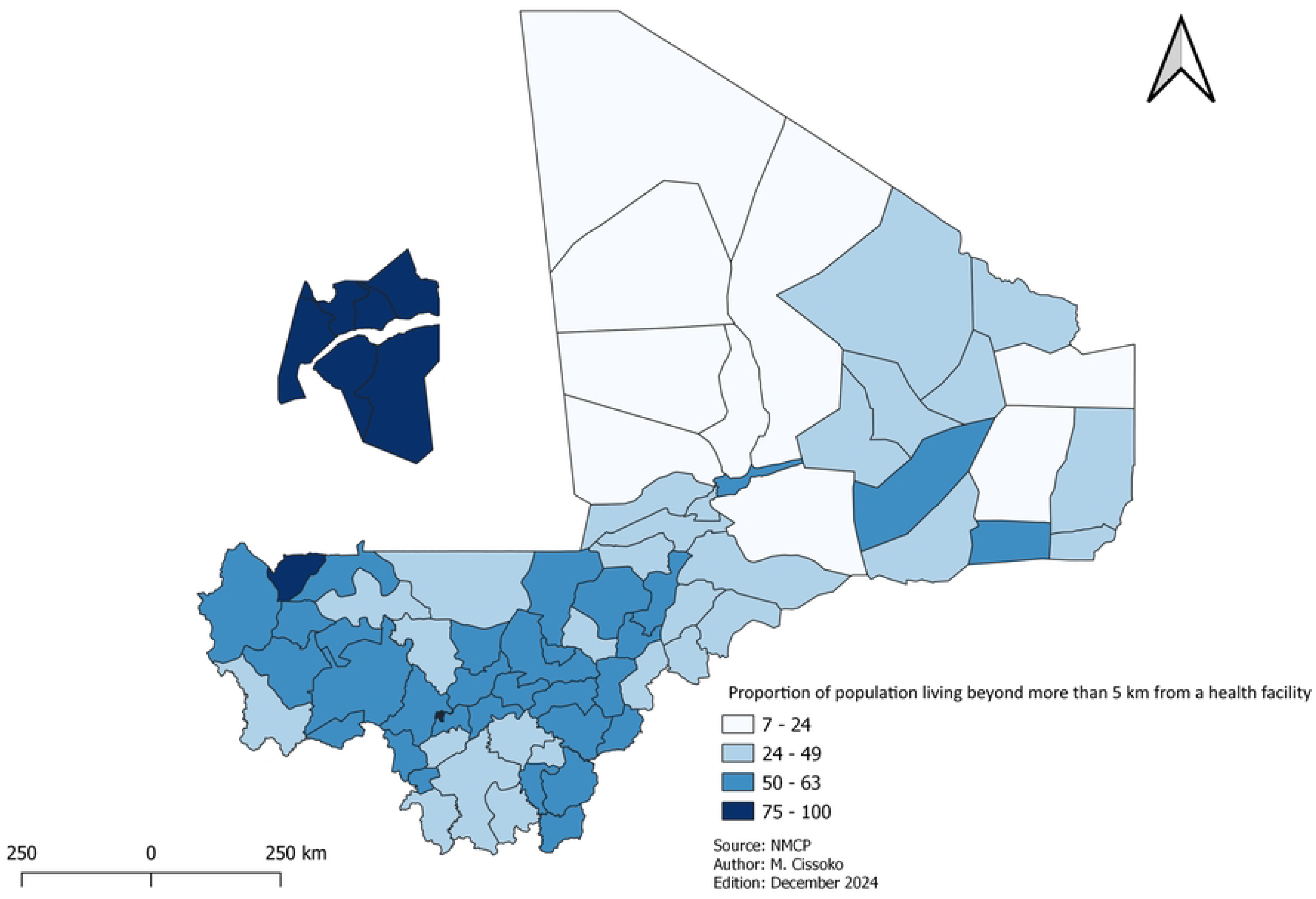
Proportion of population living beyond 5 km from a health facility.

In these underserved areas, there was an average of 5 540 people served by CHW (see Fig 2). Further, 55 of these districts were classified as High and Moderate Transmission zones.

**Fig 2.**
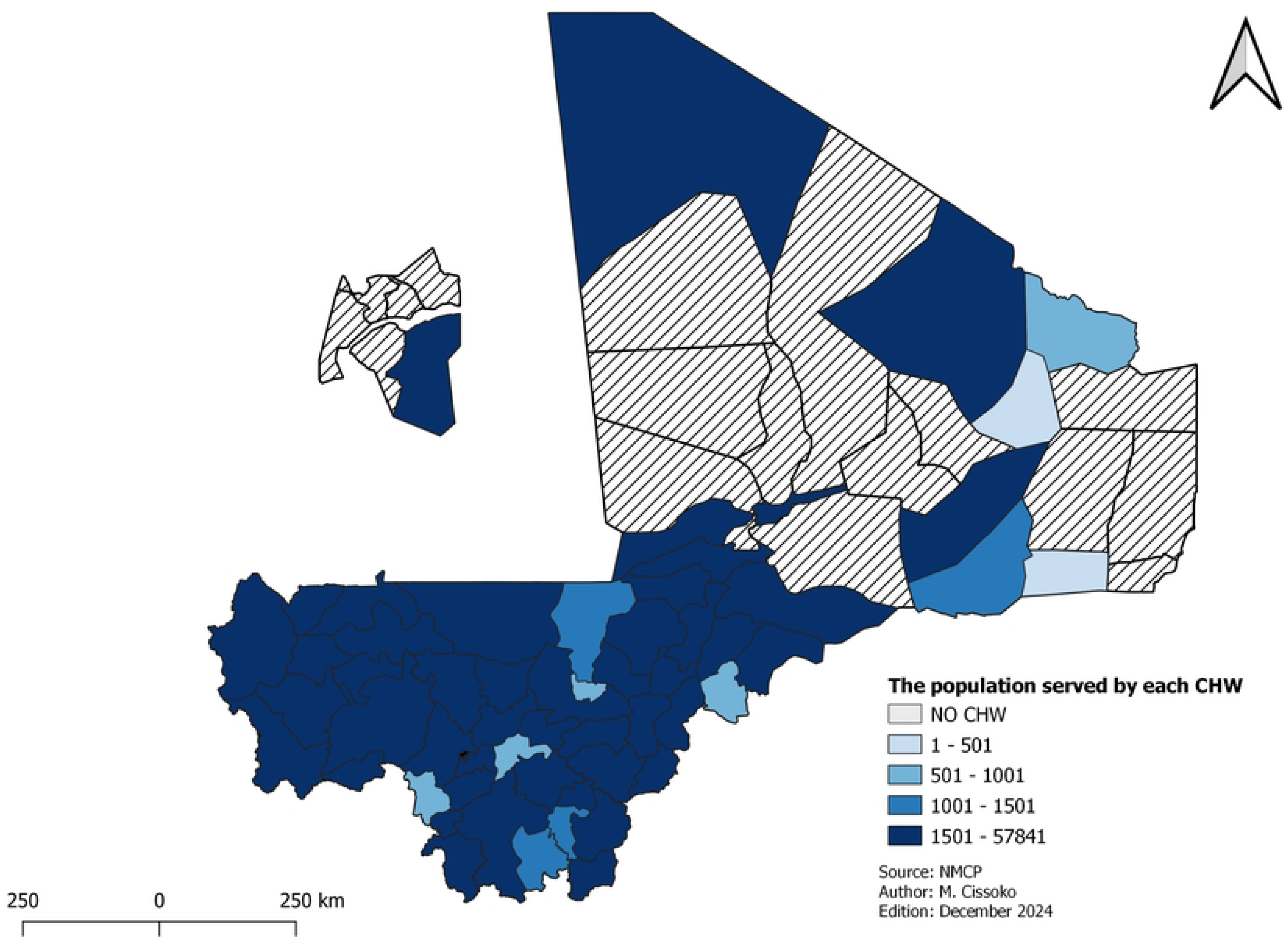
Number of populations per CHWs

Figs 1 and 2 show the population living more than 5 km from a health facility, and the number of inhabitants per CHWs.

### Rainfall

Average annual rainfall was mapped by HDs.

### Risk stratification using adjusted incidence

All HDs were classified in the transmission zone according to their adjusted incidence.

The final strata following multiple adjustments is depicted in Fig 3. About 84 % of the population lived in high and moderate strata (Table 1).

**Fig 3.**
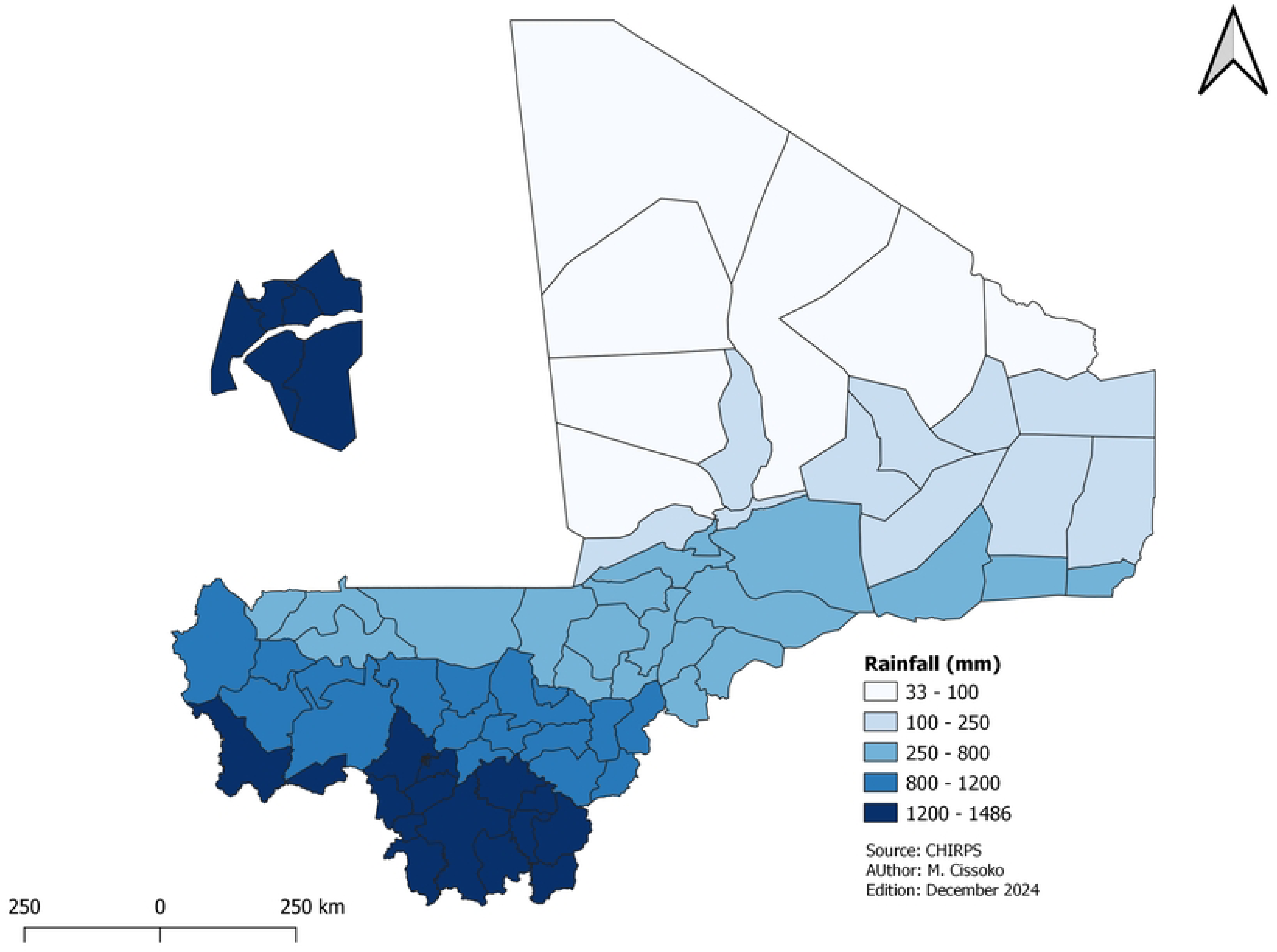
The average annual rainfall shows the variation between the southern regions (over 800mm) and the northern region (under 200mm).

**Table 1.**
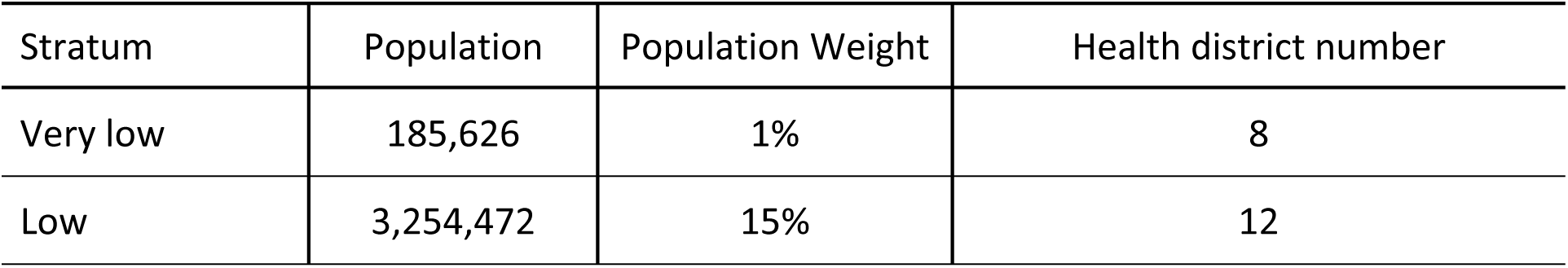
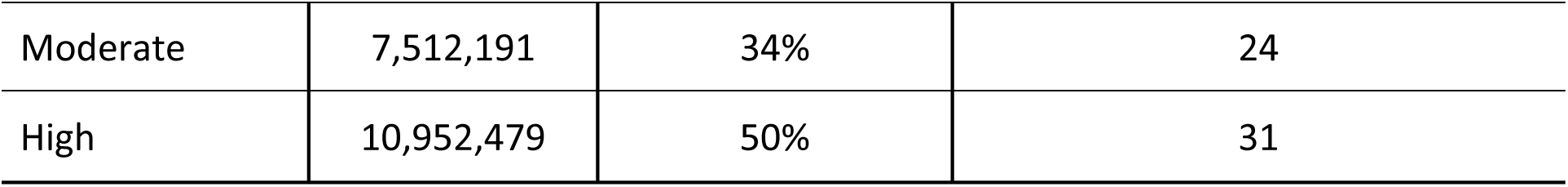
Population size by stratum.

All the health districts with moderate and high transmission were in the southern and central regions, health districts with very low transmission were in the northern regions, and health districts with low transmission were located across all regions.

### Sensitivity test

The median variation was 1.37 (CI 95%: 1.4-1.9) for the local method and 1.13 (1.1-1.5) for the WHO method. The comparison test was significant with p-value 0.02.

It is observed that the two graphs representing the variation of the methods are similar when the variations are small. The changes between the two graphs start from variations of more than 2% (Fig S1).

### Targeted malaria interventions

Considering logistical constraints and available financial resources, 13 intervention packages (Fig 3) were identified for all 75 health districts currently being implemented. The package of mix interventions tailored to each stratum is presented as follow:

#### • Malaria Seasonal Chemoprevention Prevention (SMC)

Eight health districts in the very low transmission, six health districts in Bamako (due to low malaria prevalence [24]), and four health districts with no seasonal trend over the last five years were excluded from the SMC. The Boujbeha health district (Taoudenni region) and Inekar, Anderamboucane and Tindermme (Menaka region) were non-seasonal health districts. As a result, 57 out of 75 have been selected for the SMC in Mali over the next 3 to 5 years.

By examining the monthly incidence curves, the duration of the high transmission season was determined for each year from 2019 to 2022, and the median was calculated for each health district. This median was used as the duration of the high transmission season. Based on that result, 14 health districts were qualified for five rounds, 33 for four rounds and 10 for three rounds (Fig. 4).

**Fig 4.**
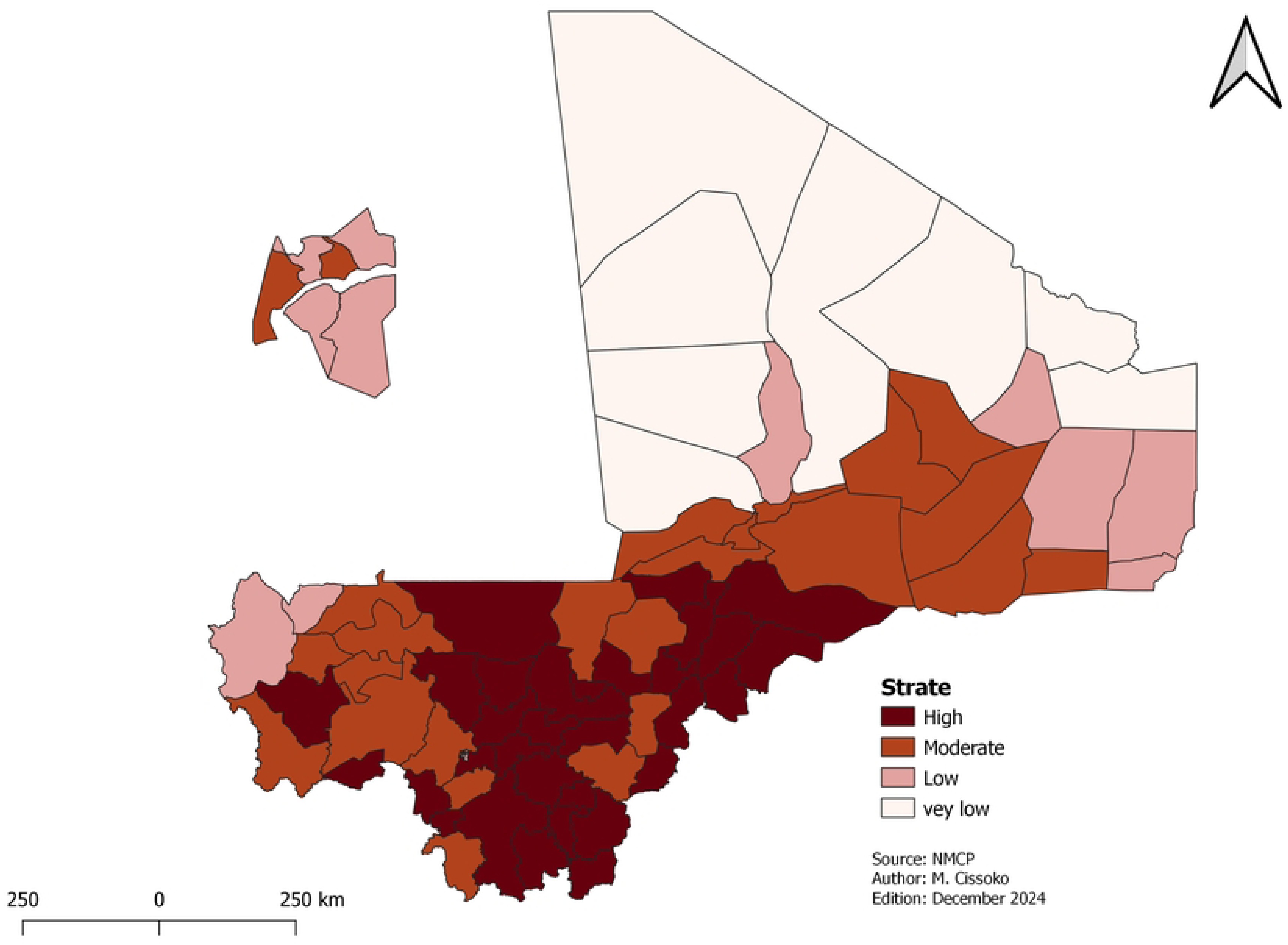
Malaria transmission strata

Fig 5 shows the number of cycles per health district and the areas without SMC. The grey health districts are not affected by this intervention.

**Fig 5.**
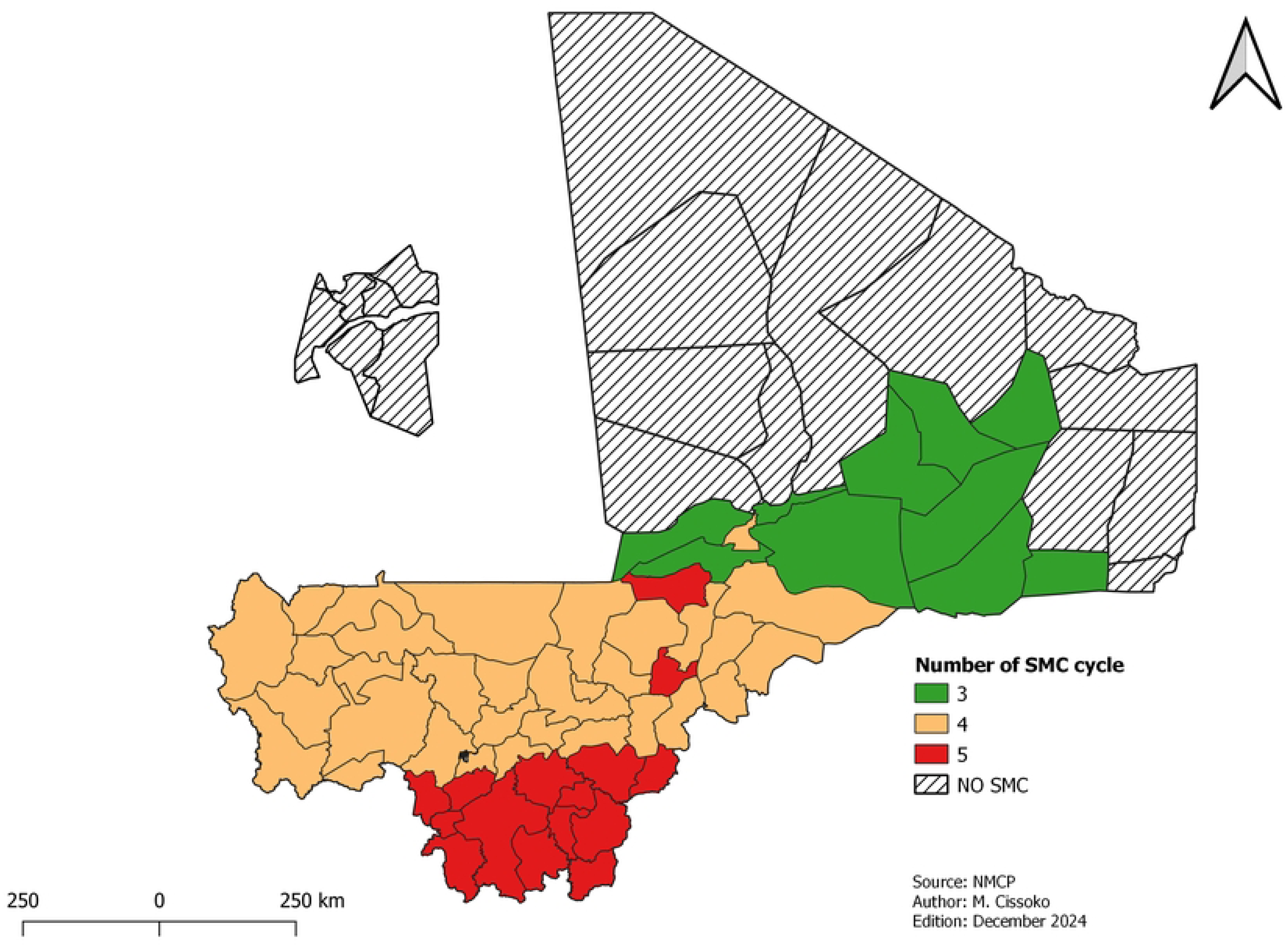
Mapping of SMC implementation and number of cycles

**Fig 6.**
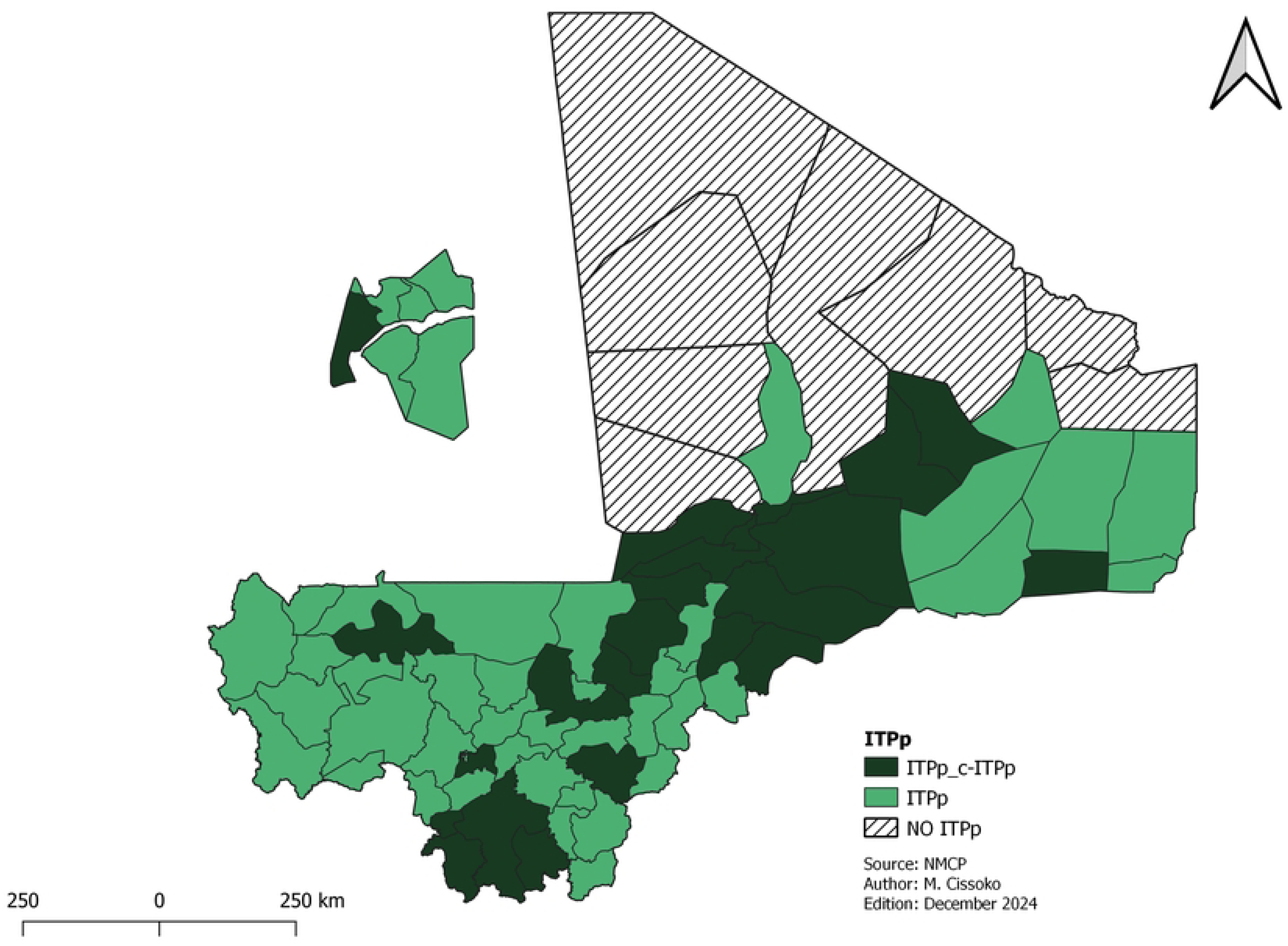
Mapping of ITPp implementation and the community ITPp levels. The grey health districts are not affected by this intervention.

**Fig 7.**
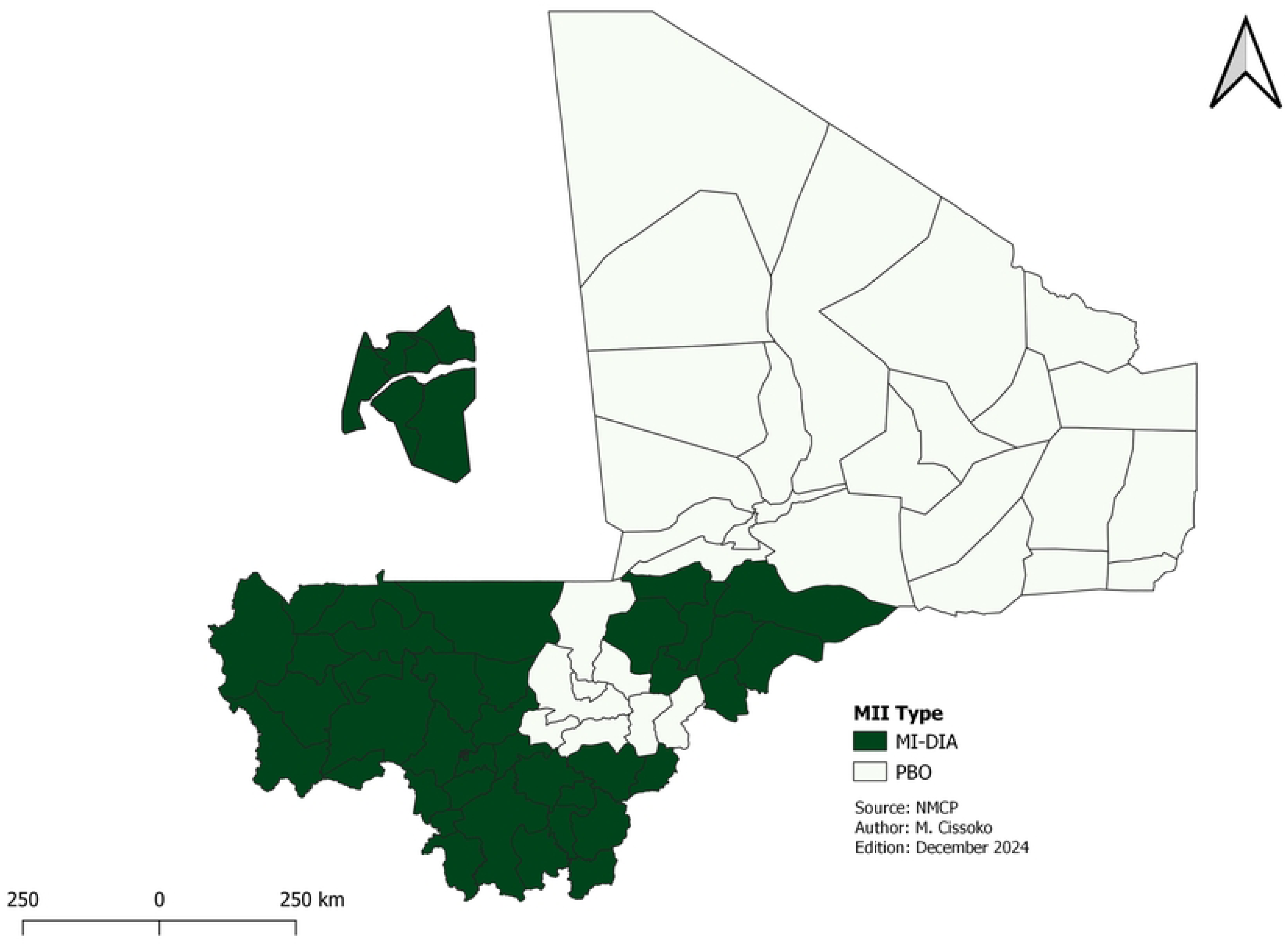
Distribution of insecticide-impregnated mosquito nets The LLINs types will be mainly new generation (Fig 7).

#### • Intermittent preventive treatment for pregnant women (IPTp)

The stratification identified 67 out of 75 health districts in areas of low, moderate, and high transmission areas eligible for IPTp. All the eight health districts excluded from this intervention were in two northern regions of the country, where malaria transmission was very low. This represented 22 health districts. Its implementation began with a pilot phase in 2 health districts in the Ségou region (San and Bla) which have good CHW coverage.

#### • Vaccination against malaria

Based on the stratification, 19 health districts are initially selected to receive the malaria vaccine in 2025 due to limited availability of the vaccine. An additional 36 health districts will receive vaccine in 2026. All these districts were in high and moderate malaria transmission with high mortality.

#### • Distribution of long-lasting insecticidal net (LLINs)

We identified 61 out of 75 health districts with high, moderate, and low transmission eligible for the mass distribution campaign versus 68 HDs for the first stratification which identified six health districts in Bamako (low prevalence of malaria) to be excluded. Additionally, 8 health districts with very low transmission were again excluded from the mass campaign plan in 2026. The LLINs mass campaign will be conducted in five regions of the southern and central parts of the country: Kayes, Koulikoro, Sikasso, Ségou and Mopti. Due to security concerns, community leaders, with support from non-governmental organizations (NGO), will take the leadership for LLINs community distribution in the northern regions: Timbuktu, Gao, Kidal, Ménaka and Taoudenni during the mass campaign.

To maximize coverage and ensure that vulnerable populations receive essential protection against malaria, LLINs distribution will also be organized for teachers and students in basic schools in Bamako, as well as for vulnerable groups (displaced, orphans, returnees and the prisoners) in districts not included in the campaign. This distribution will take place during the years of the mass distribution campaign. This specific distribution will help improve the fight against malaria in Bamako.

All health districts in the country, regardless of stratum, will continue to receive LLINs through routine distribution during the immunization against measles for children under 1 year and for pregnant women starting at their first prenatal visit.

In addition to stratification risk results, we considered insecticide resistance data and the concerns over the withdrawal of indoor residual spraying (IRS) from three health districts in the Mopti region to identify the most suitable of LLINs for each district. In total, 38 health districts in the Kayes, Koulikoro, Sikasso and Mopti regions will receive dual active ingredient (dual AI) LLINs. These districts accounted for 80% (25 out of 31) of health districts with high transmission and 44% (11 out of 25) of health districts with moderate transmission. Although most health districts in the Ségou region fall within moderate to high transmission areas, and recent data from sentinel sites suggesting effectiveness of PBO LLINs in other parts of the country [9], all districts of this region will continue to receive these nets through the campaign and the routine distributions. School distribution using dual AI LLINs with pyrethroid resistance and low susceptibility to LLINs-PBOs in 2021, according to the PNLP’s entomological surveillance report on sentinel sites.

Similarly, 10 health districts with moderate transmission and 3 health districts with low transmission in the Tombouctou, Gao, Kidal, Taoudenni and Menaka regions, where resistance to PBO LLINs has not been documented, will also receive PBO LLINs these.

#### • Indoor Residual Spraying (IRS)

IRS has been proposed for the response to the malaria epidemic.

#### • Malaria biological diagnostic tests

Malaria diagnostic tests will be performed using HRP2/3 rapid diagnostic tests (RDTs) and microscopy in all regions, except for Ménaka and Kidal, where the prevalence of *P. vivax* is 21% and 4%, respectively. In these regions, the new malaria national strategic recommends using Pf/Pv RDTs for rapid diagnosis. Currently, there was no alert on HRP2/3 deletion in Mali. The national malaria control program will continue to investigate possible deletion and the distribution of plasmodial species across the country and will adjust the use of RDTs based on the results.

#### • Antimalaria treatment

The treatment of malaria will continue to be done with ACT for uncomplicated malaria and injectable artesunate for complicated cases, regardless of the level of malaria transmission.

CHWs will keep administrating artemether-lumefantrine (AL) at community level. Starting in 2025, dihydroartemisinin-piperaquine (DHA-PPQ) will be introduced at health facilities, beginning in Mopti region, with plan to extend to Sikasso and Koulikoro region in the future, as these areas had the highest parasite prevalence’s, and were in the high transmission strata.

In areas with very low transmission, as well as in Kidal and Taoudenni, patients with *P. falciparum malaria* will receive a single dose of Primaquine alongside ACT to prevent relapses by eliminating gametocytes from the blood. Pregnant women, children under 6 months and breastfeeding women are not eligible for Primaquine.

Due to lack of systematic G6PD deficiency screening in Mali, radical treatment of *P. vivax* will not begin until the second year in 2026. The use of Pf/Pv RDTs will make it possible to estimate the number of patients infected by *P. vivax*. The rapid G6PD test will then be used in the Kidal and Ménaka regions to detect its deficiency and determine the possibility of radical treatment with primaquine. Based on trends observed, the program may consider introducing rapid G6PD deficiency tests, along with high dose primaquine treatment for radical cure.

Fig 8 shows the case management guidelines according to parasite species, the objective of elimination in areas of very low transmission and the multi-first-line therapeutic combinations.

**Fig 8.**
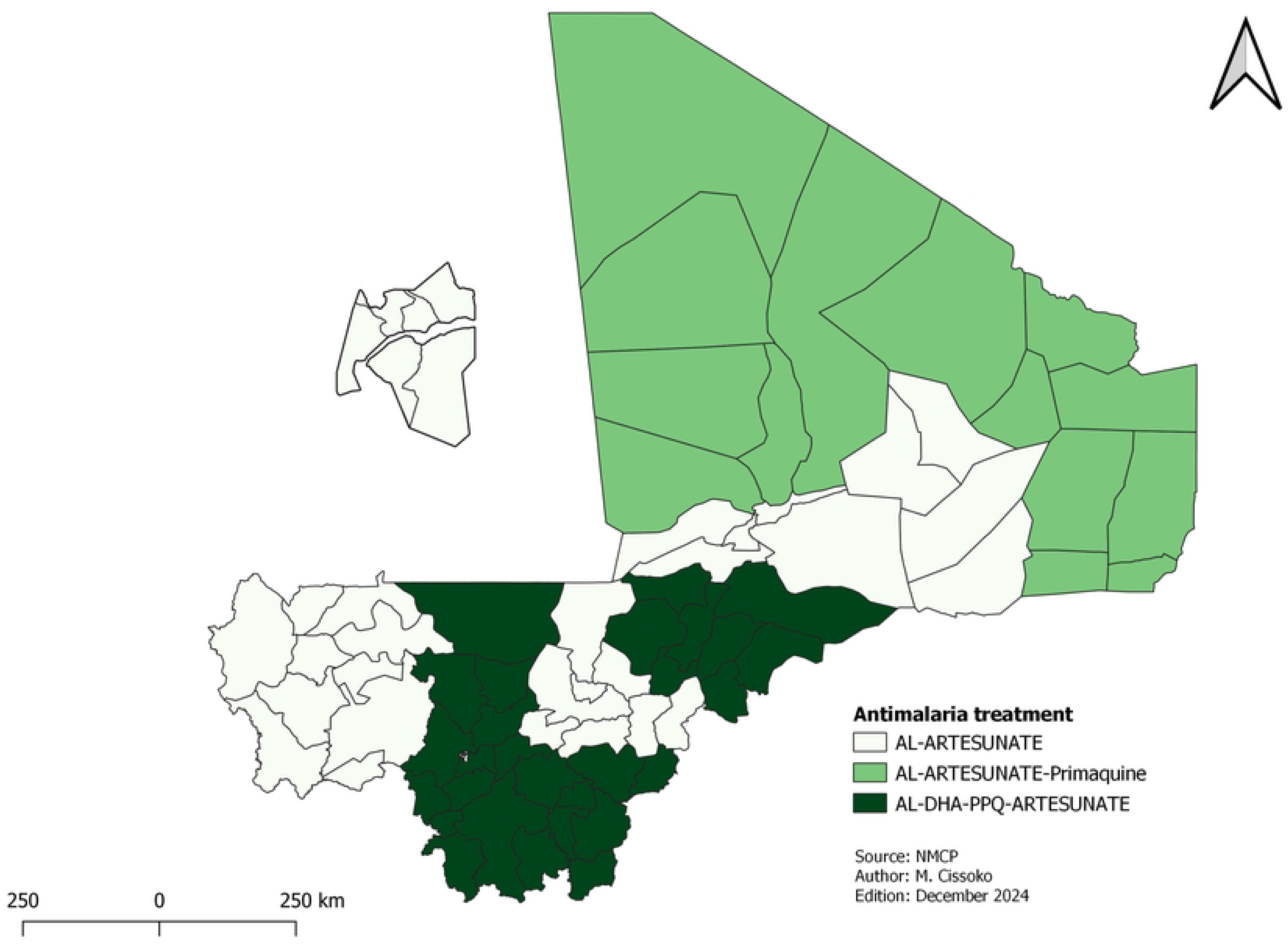
Antimalaria treatment

#### • Malaria surveillance

Routine surveillance will be maintained on weekly and monthly basis across all health districts in low, moderate and high transmission zones, while case-by-case surveillance will be conducted in areas with very low transmission.

Entomological surveillance will be conducted at selected sentinel sites level across all strata. To monitor *Anopheles stephensi*, airports, bus stations and border areas will be targeted.

Finally, 19 intervention packages were selected for the 75 health districts over the next 3 to 5 years.

Table 2 and Fig 9 summarize the interventions selected by the health district. The health districts with the identified interventions are grouped together and are the same color.

**Fig 9.**
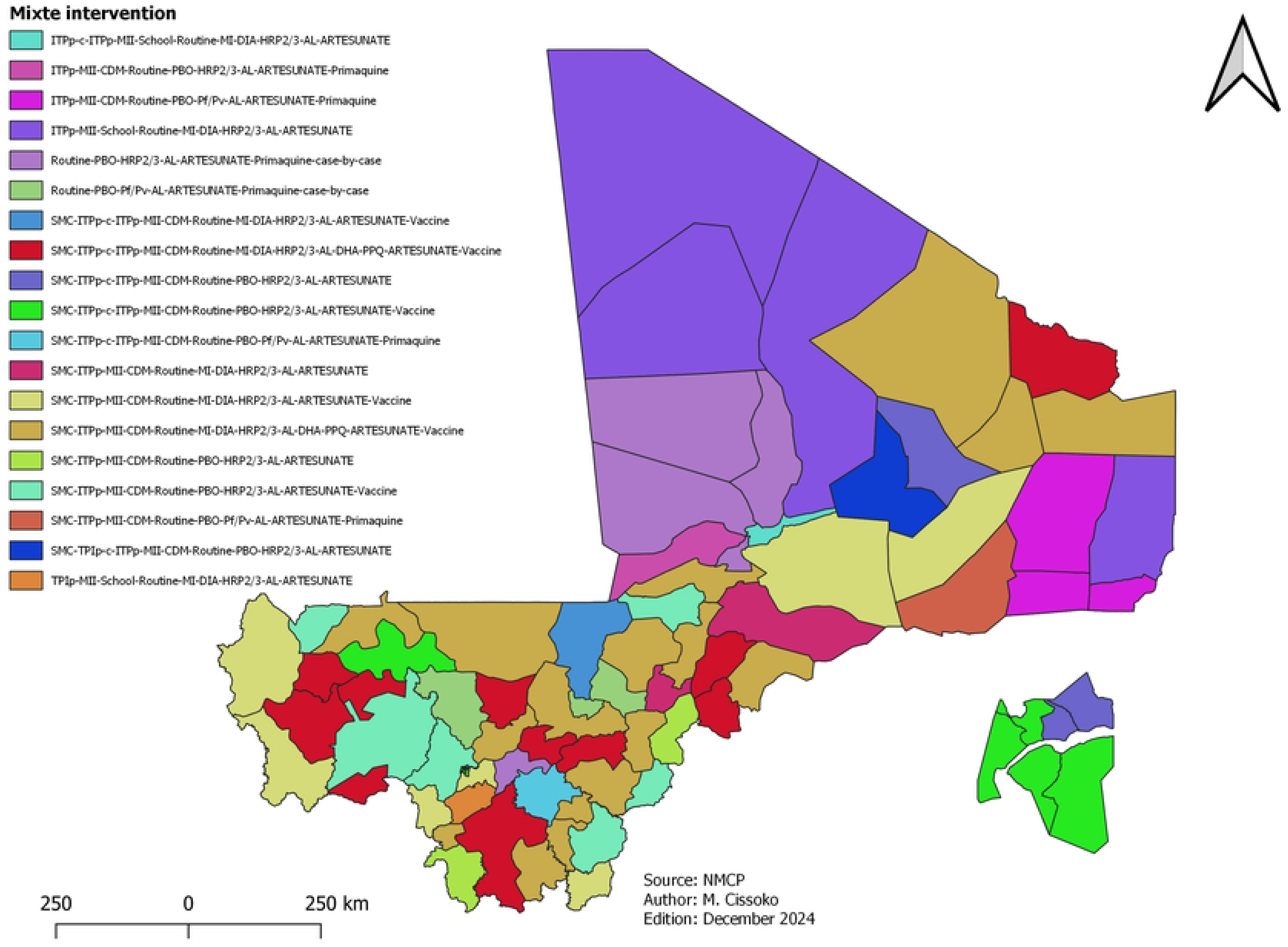
Map of the malaria control intervention mix in Mali.

**Table 2.** The number of health districts per intervention packages.

| Packages | Number |
| --- | --- |
| ITPp-c-ITPp-LLIN-School-Routine-MI-DIA-HRP2/3-AL-ARTESUNATE | 1 |
| ITPp-LLIN -CDM-Routine-PBO-HRP2/3-AL-ARTESUNATE-Primaquine | 1 |
| ITPp- LLIN -CDM-Routine-PBO-Pf/Pv-AL-ARTESUNATE-Primaquine | 3 |
| ITPp-LLIN -School-Routine-MI-DIA-HRP2/3-AL-ARTESUNATE | 4 |
| Routine-PBO-HRP2/3-AL-ARTESUNATE-Primaquine-case-by-case | 5 |
| Routine-PBO-Pf/Pv-AL-ARTESUNATE-Primaquine-case-by-case | 3 |
| SMC-ITPp-c-ITPp-LLIN -CDM-Routine-MI-DIA-HRP2/3-AL-ARTESUNATE-Vaccine | 1 |
| SMC-ITPp-c-ITPp-LLIN -CDM-Routine-MI-DIA-HRP2/3-AL-DHA-PPQ-ARTESUNATE-Vaccine | 11 |
| SMC-ITPp-c-ITPp-LLIN -CDM-Routine-PBO-HRP2/3-AL-ARTESUNATE | 3 |
| SMC-ITPp-c-ITPp-LLIN -CDM-Routine-PBO-HRP2/3-AL-ARTESUNATE-Vaccine | 5 |
| SMC-ITPp-c-ITPp-LLIN -CDM-Routine-PBO-Pf/Pv-AL-ARTESUNATE-Primaquine | 1 |
| SMC-ITPp- LLIN-CDM-Routine-MI-DIA-HRP2/3-AL-ARTESUNATE | 2 |
| SMC-ITPp- LLIN-CDM-Routine-MI-DIA-HRP2/3-AL-ARTESUNATE-Vaccine | 7 |
| SMC-ITPp- LLIN-CDM-Routine-MI-DIA-HRP2/3-AL-DHA-PPQ-ARTESUNATE-Vaccine | 17 |
| SMC-ITPp- LLIN-CDM-Routine-PBO-HRP2/3-AL-ARTESUNATE | 2 |
| SMC-ITPp- LLIN-CDM-Routine-PBO-HRP2/3-AL-ARTESUNATE-Vaccine | 6 |
| SMC-ITPp- LLIN-CDM-Routine-PBO-Pf/Pv-AL-ARTESUNATE-Primaquine | 1 |
| SMC-TPIp-c-ITPp-LLIN-CDM-Routine-PBO-HRP2/3-AL-ARTESUNATE | 1 |
| TPIp- LLIN -School-Routine-MI-DIA-HRP2/3-AL-ARTESUNATE | 1 |

## Discussion

This research was focused on the role of malaria risk stratification in targeted interventions for malaria control. Stratification of malaria risk showed that most health districts were at the high area transmission, i.e. around 80 % of the population. Malaria diagnosis and treatment have been adapted with the introduction of DQA-PPQ and rapid *Pf/Pv*-based RDTs. As for LLINs, the mass campaign will cover only the 61 health districts at high risk of transmission. Based on the eligibility criteria, 18 HDs were excluded from the SMC, compared with 17 in the first stratification. Other risk factors such as vector resistance to insecticides and limited financial funding were considered in the LLINs distribution.

Analysis of geographical accessibility indicates that over half of the health districts had a large population living beyond 5 km from a health facility (see Fig. 1). These health districts have been prioritized for the deployment of CHWs to improve access to malaria management services. According to the national plan for essential care in the community, the standards are 100 to 500 inhabitants in the northern districts and 500 to 1,000 inhabitants in the south per CHW. The deployment of CHWs was intended to improve the management of malaria cases [25]. However, the number of CHWs remained below standard in the southern health districts, which account for 80% of malaria cases. The number of CHWs needs to be increased in these health districts to ensure rapid screening and treatment, and behavioral change in the use of preventive measures such as LLINs. Community health care faces other problems, including the regular payment of the CHWs motivation, and the stability of community health workers at their sites [26].

For a better estimate of the number of cases and incidence, the study considered the adjustment of factors such as reporting, the rate at which the diagnostic test was carried out and the prevalence of fever cases receiving health care. The data used concerned patients who visited the health facilities and whose data had been correctly reported. The various adjustments made it possible to take these shortcomings into account. In this study, non-use of care was estimated based on rate of healthcare use in health facilities and care-seeking rate, which was available for all health districts, unlike the prevalence of fever cases having care, which was at regional level [23]. Health districts in the same region do not have similar levels of access to and use of services. The WHO method proposes the preventive use of healthcare. This information was available at regional level in Mali. We therefore used the attendance rate, which was available for all health districts. Although this method is recommended for correcting routine data[27]. Comparison of the variation between the two models showed similarities except for a few health districts. These results suggest that the two methods can be used, especially if the DHS data are old, as in Mali, where the last survey was conducted in 2018.

The stratification of Mali in 2022 identified 57 health districts eligible for the SMC versus 59 in the first stratification. Of these, one health district in the Ménaka region was excluded because of the non-seasonality trend. This situation is due linked to the displacement of populations due to the security crisis. The large city of Bamako, which is an urban area with 6 communes, was excluded from the SMC campaign based on the malaria prevalence malaria. However, specific stratification at the health area level is necessary to identify certain eligible sites for the implementation of SMC, such as peri-urban quarters and internally displaced persons sites. According to modeling from 2021 [28], achieving 80% coverage in four rounds will have a greater impact than five rounds if the coverage is inadequate. The NMCP in collaboration with its partners, is ramping up efforts to improve coverage and maximize the effectiveness of SMC. These initiatives include evaluating the entire SMC process, involving the community in the monitoring of drug administrations, and integrating SMC with other activities such as fever screening, screening for malnutrition and prenatal care services [28].

IPTp coverage was still below the national target. An innovation in ITPp has been its introduction at community level. Dispensing with community health workers removes the barrier from the 2^nd^ dose uptake, as the pregnant woman can receive subsequent doses at home through the community health worker. This community-based approach has produced very good results in several countries. It is acceptable to communities and CHWs [29,30]. But during the implementation of the community IPTp in Mali, criteria used were challenging due to insecurity, low number of CHWs (i.e. HDs did not have enough CHWs to have an impact), and payment of CHWs motivation. For example, HDs without CHWs or with insufficient CHWs were excluded. The NMCP has adjusted the implementation to ensure that the intervention has a strong impact by covering the whole Mopti region, which has the highest malaria prevalence in the country.

The use of LLINs PBOs compared with DUALs is explained by a lack of funding. But only the Ségou region in the south, which has no metabolic resistance, will continue to use PBO LLINs. Health districts in the north, where there was no evidence of pyrethroid resistance in the major vector, will distribute PBOs instead of standard LLINs. Standard LLINs have not been included in the current strategic plan because of widespread resistance to pyrethroids. Distribution methods have been diversified with school [31] and community distribution to increase the universal coverage rate, which remains low at 44%. Diversifying distribution strategies can help to reduce the risk of malaria. After the LLINs distribution campaign in 2023, the number of malaria cases fell by 10%, something that had not happened since 2017 [3]. Weaknesses in routine distribution, such as stock-outs, must be considered. The same applies to access to health facilities, where co-payment is required for consultations.

The first stratification proposed HRP3/3 RDTs for the whole country, because *P. falciparum* was the majority species. Based on recent studies, a variation was observed in certain areas. In terms of management, the improvements diagnosis, the *Pf/Pv* RDTs are introduced in two regions to diagnose malaria cases of *P. vivax*.

For the treatment improvements, a second artemisinin-based combination therapy had been integrity the quantification for 2025. Artemether-Lumefantrine has been used in Mali for more than 10 years. The introduction of DHA-PPQ will contribute to delaying the resistance to Artemether-Lumefantrine. DHA-PPQ will be introduced gradually. This delay will allow the NMCP to revise its monitoring and evaluation tools so that it can report the proportion of each ACT. The introduction of DHA-PPQ is also justified by the shortcomings in health care provider compliance with guidelines of uncomplicated malaria treatment, which may contribute to increasing the lack of confidence since many cases of uncomplicated malaria were treated with injectable forms.

Primaquine is not currently included in the national guidelines because of the recommendation to stop monotherapy. It is included in the guidelines and used to prevent *P. falciparum* cases for the HDs of very low transmission and the radical cure of *P. vivax* cases. However, primaquine should not be used for *P. vivax* malaria case treatment soon because of the risk of hemolysis in patients with G6PD deficiency. Due to these conditions, the NMCP will introduce rapid G6PD screening tests in these regions from 2026. Incidence data show that the Menaka region has moderate transmission, which may explain the multiple episodes. For pregnant women who will not be treated with primaquine, the alternative will be the use of ACT only.

The proposed methodology, based on a qualitative approach, differs from that proposed in other studies. The methodology proposed by our study is based on relevant data from interventions to define deployment zones. Some studies have used mathematical methods, using socio-economic or meteorological factors to target interventions [32,33].

This research presents some strengths and innovative methodologies. The data from DHIS2, which is the platform for routine data reporting, contributed to improving this stratification. In addition, we adjusted malaria cases to better estimate the risk of malaria. For the targeting of malaria control interventions, changes have been made to achieve an impact on malaria transmission. It needs to be updated to consider malaria cases by parasite species, the type of RDT used, treatment with DHA-PPQ, primaquine, malaria vaccination and c-IPTp. To make these innovations successful, a series of skills-building training sessions will be needed for health service providers in Mali. Normative documents will also be revised for these training sessions in 2024.

The main limitations of the study are the failure to take account of meteorological data in determining malaria risk and the classification by stratum. These data have an impact on malaria transmission in local contexts such as flood zones. The second limitation of this study may be the use of malaria prevalence to target certain interventions in the city of Bamako. This suggests that future stratification should consider this particularity of Bamako by showing peri-urban areas where the risk of malaria is high. In Mali, studies have already shown the link between malaria transmission and meteorological factors. Based on this information, this simplified study of malaria risk stratification could become a model for other countries.

## Conclusion

The malaria risk assessment allowed us to define the different strata of transmission. Analysis of the situation revealed a need to improve interventions’ delivery. Stratification has made it possible to target interventions according to objective criteria, and to propose innovations such as the introduction of a second first-line ACT for the treatment of uncomplicated malaria, the use of new RDTs to confirm cases of *P. vivax* in certain regions, and the introduction of Primaquine in certain regions. Carrying out this exercise periodically and regularly will enable the mixed map of malaria control interventions to be adapted for optimum use of resources.

## Data Availability

Data can be shared on request, with reasons.

## Acknowledgments

Our thanks go to the participants of the analysis workshop in the regional health departments of Kayes, Sikasso, Ségou and Mopti.

## Author contributions

Data was collected and cleaned by MC and IAC. The analysis plan was drawn up by MC and approved by IS, ST and JG. Analyses were performed by MC, MK, MM, VS and IS. The manuscript was redacted by MC, SC, TB, SD, AK and SF. The manuscript was read and approved by all authors.

---

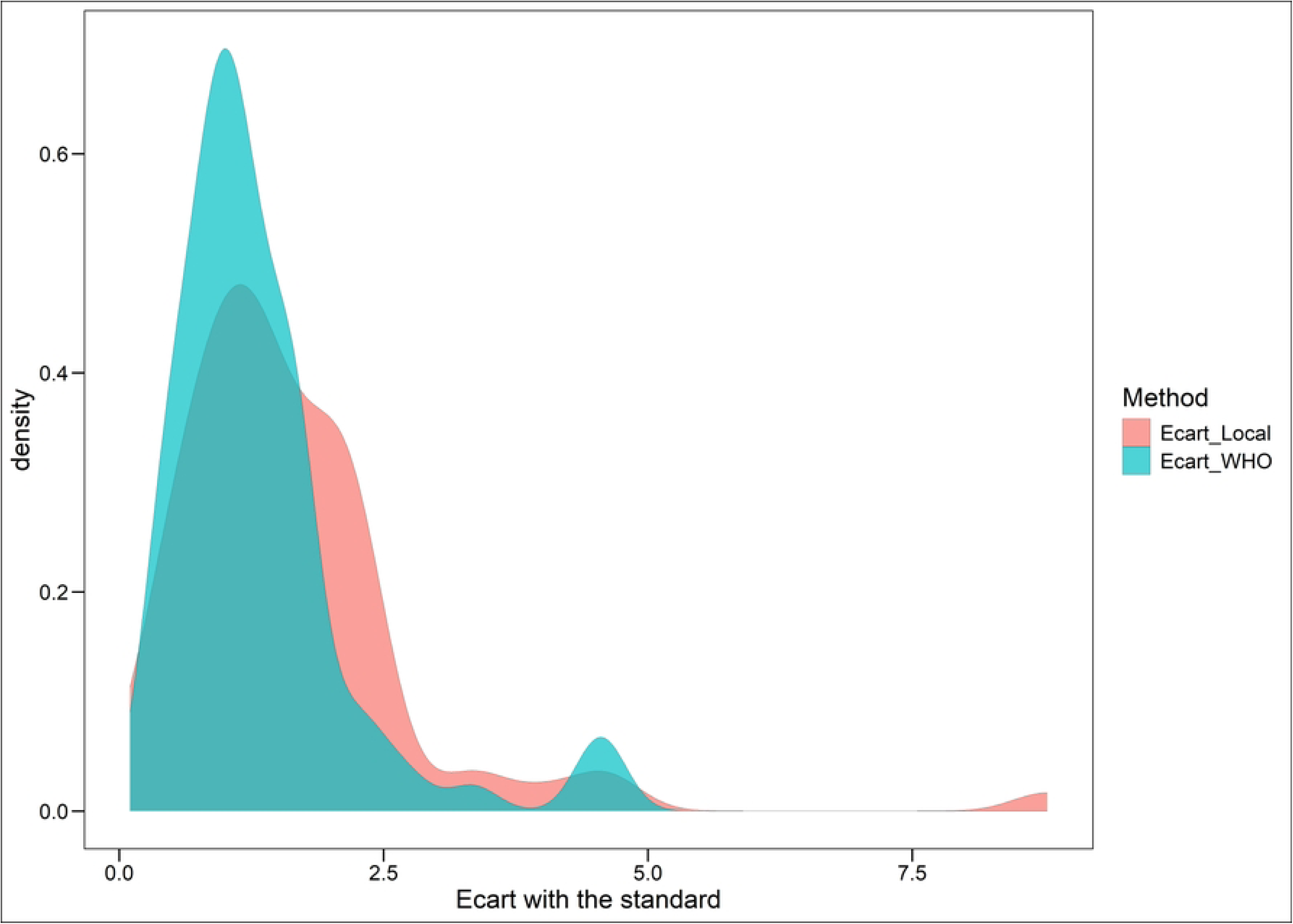

